# Maternal and perinatal outcomes among pregnant women with sickle cell disease in Uganda: a prospective cohort study

**DOI:** 10.64898/2026.07.02.26357097

**Authors:** Jackline Akello, Sarah Kiguli, Grace Ndeezi, Musa Sekikubo, Savio Mwaka, Ian Munabi, Ruth Namazzi, Deogratias Munube, David Mukunya, Pebalo Francis, Kenneth Mugabe, Joseph Rujumba, Annettee Nakimuli

**Affiliations:** Department of Obstetrics and Gynaecology, School of Medicine, Makerere University, College of Health Sciences, Kampala, Uganda; Department of Pediatrics and Child Health, School of Medicine, Makerere University, College of Health Sciences, Kampala; Department of Anatomy, School of Biomedical Sciences, College of Health Sciences, Makerere University, Kampala; Department of Public Health, Busitema University, Mbale; Department of Obstetrics and Gynaecology, School of Medicine, Gulu University, Gulu; Department of Obstetrics and Gynaecology, Mbale Regional Referral Hospital, Mbale; Research Department, Infectious Diseases Research Collaboration, Kampala, Uganda; Research and Innovations Department, Uganda Institute of Information and Communication Technology, Kampala, Uganda

**Keywords:** sickle cell disease, pregnancy, maternal complications, perinatal outcomes, Uganda, Africa, prospective cohort

## Abstract

**Background:** Pregnancy in women with sickle cell disease (SCD) is associated with substantial maternal and perinatal risk, but prospective data from Uganda remain limited. We described the clinical profile, maternal complications, and fetal and neonatal outcomes among pregnant women with confirmed SCD receiving tertiary-level care in Uganda.

**Methods:** We conducted a hospital-based prospective cohort study of 159 pregnant women with confirmed HbSS at Kawempe National Referral Hospital and Mbale Regional Referral Hospital, Uganda, between October 2024 and March 2026. Participants were recruited consecutively during antenatal care or at admission for delivery and followed until maternal discharge. Outcomes were summarized descriptively using explicit denominators. Maternal complications were reported at woman level, while fetal and neonatal outcomes were reported at fetus/newborn level to account for multiple gestations. Secondary regression analyses were treated as exploratory.

**Results:** The mean maternal age was 24.3 years (SD 4.7), and 82/159 women (51.6%) were referred in labor or on admission rather than being known from the study-site antenatal clinics. At baseline, median hemoglobin was 7.5 g/dL (IQR 6.4–8.75), and 118/132 women (89.4%) had hemoglobin below 10 g/dL. The most frequent maternal complications were anemia (93/159, 58.5%), vaso-occlusive crisis (87/159, 54.7%), acute chest syndrome (31/159, 19.5%), recurrent infections (27/159, 17.0%), postpartum hemorrhage (21/159, 13.2%), and preeclampsia (17/159, 10.7%). Maternal death occurred in 15/159 women (9.4%). Among women with recorded mode of delivery, 107/137 (78.1%) delivered by caesarean section. The 159 women contributed 166 fetal outcomes, including seven twin pregnancies. Of these, 22/166 (13.3%) were abortions. Among 144 fetal/newborn outcomes after exclusion of abortions, 119/144 (82.6%) were live births and 25/144 (17.4%) were stillbirths. Early neonatal death occurred in 12/119 live births (10.1%). Low birthweight occurred in 76/117 newborns with recorded birthweight (65.0%), and NICU admission was recorded in 83/118 newborns with recorded admission status (70.3%).

**Conclusion:** Pregnancy among women with SCD receiving tertiary referral care in Uganda was characterized by high maternal morbidity, substantial maternal mortality, and poor fetal and neonatal outcomes. The findings support early identification and referral, structured multidisciplinary antenatal care, reliable transfusion readiness, rapid escalation for acute chest syndrome, preeclampsia and infection, and close coordination between obstetric and neonatal services.

## Introduction

Sickle cell disease (SCD) is among the most common inherited haemoglobin disorders worldwide and remains a major cause of morbidity and premature mortality, particularly in sub-Saharan Africa (1–3) . Existing evidence from Ghana, Tanzania, Jamaica, the UK, and other settings consistently describes SCD pregnancy as high risk, with severe anaemia, vaso-occlusive crises, infection, pulmonary complications, and thromboembolic events contributing importantly to poor outcomes(4–7) . Uganda carries a high SCD burden, with national surveillance data showing substantial geographic variation in sickle cell trait and disease and suggesting that approximately 15,000 babies are born annually with SCD (8) . As survival into adolescence and adulthood improves, increasing numbers of women with SCD are reaching reproductive age, making pregnancy-related care an urgent clinical and public-health priority.

Pregnancy superimposes haemodynamic, inflammatory, and thrombotic stress on an already vasculopathic and haemolytic disorder. Women with SCD are therefore at increased risk of painful vaso-occlusive episodes, severe anaemia, acute chest syndrome, thromboembolism, infection, hypertensive disorders of pregnancy, haemorrhage, preterm birth, fetal growth restriction, stillbirth, and maternal death. Meta-analyses and cohort studies have consistently shown that these risks remain elevated across both low-income and high-income settings, although the burden is generally greatest where access to specialist care, transfusion support, and critical care is limited (3,6,7,9).

Despite Uganda’s high SCD burden, prospective pregnancy-specific data remain limited, particularly data that separate maternal, fetal, and neonatal denominators and describe outcomes within referral-hospital care pathways. Such data are needed to inform context-appropriate models of antenatal surveillance, referral, delivery planning, transfusion readiness, and neonatal care for women with SCD in high-burden African settings. We therefore aimed to describe the clinical profile, maternal complications, and fetal and neonatal outcomes among pregnant women with confirmed HbSS receiving tertiary-level care in Uganda.

## Methods

We conducted a hospital-based prospective cohort study of pregnant women with confirmed sickle cell disease receiving care at Kawempe National Referral Hospital and Mbale Regional Referral Hospital, Uganda, between October 2024 and March 2026. The study was designed primarily to describe maternal complications and pregnancy, fetal and neonatal outcomes in referral-hospital practice. Exploratory regression analyses were conducted secondarily to identify clinical factors associated with selected adverse outcomes and should be interpreted as hypothesis-generating.

### Study participants

The study population comprised pregnant women with confirmed HbSS receiving care at the two study hospitals during the study period. Women were eligible if they had confirmed HbSS, were pregnant at enrolment, and provided written informed consent. Women without genotype confirmation were not enrolled. Women with no retrievable pregnancy, delivery, or discharge outcome information after review of available facility records and follow-up attempts were excluded from the final analytic dataset.

### Recruitment and follow-up

Participants were recruited consecutively during antenatal care or at admission for delivery. At enrolment, baseline sociodemographic, obstetric and clinical information was collected using structured study forms and facility records. For women enrolled before delivery, follow-up information was updated during routine antenatal contacts where applicable, interim admissions, the delivery admission and the immediate post-delivery hospital stay. Maternal outcomes were ascertained until maternal discharge. Newborn outcomes were abstracted from delivery, neonatal-unit and discharge records where available. There was no scheduled post-discharge follow-up for this analysis. Participant flow and analytic denominators were summarized using a STROBE-style approach. The numbers screened, eligible, consented, enrolled, excluded, and included in the final mother-level analytic cohort were recorded. Denominators for delivery, fetal, and neonatal outcomes were defined according to availability of outcome-specific information, and are reported explicitly in the Results tables.

### Confirmation of sickle cell disease

Sickle cell genotype was confirmed for all enrolled participants by haemoglobin electrophoresis performed at the Central Public Health Laboratories, Uganda’s national reference laboratory. Only women with confirmed HbSS were included in the study.

### Data collection and variables

Data were collected electronically using a structured data-collection platform. Baseline variables included sociodemographic characteristics, obstetric history, admission status, haemoglobin concentration, HIV serostatus, blood group, renal and liver function test results, transfusion history, hydroxyurea exposure, antenatal medications, number of hospital admissions during pregnancy, antenatal care attendance, and psychosocial support indicators.

Maternal complications assessed during pregnancy included anaemia, vaso-occlusive crisis, acute chest syndrome, recurrent infections, postpartum haemorrhage, preeclampsia, deep vein thrombosis, malaria, antepartum haemorrhage, pulmonary embolism, acute kidney injury, chorioamnionitis, severe oligohydramnios, surgical-site infection or puerperal sepsis, and intrauterine fetal death. Maternal outcomes included HDU or ICU admission, maternal death, mode of delivery, indications for caesarean section, and post-delivery length of stay. Neonatal outcomes included live birth, stillbirth, abortion, early neonatal death, APGAR score, birthweight, NICU admission, indications for NICU admission, and NICU length of stay.

Repeated hospital admissions during pregnancy were not entered as new participant records. Instead, events and reasons for admission were recorded within each participant’s existing study record.

### Operational definitions of clinical variables

The following operational definitions were used to harmonise clinical variables across study forms and routine clinical records. Anaemia was defined as haemoglobin concentration below 10 g/dL during pregnancy or delivery admission. Severe anaemia was defined as haemoglobin concentration below 7 g/dL. Vaso-occlusive crisis was defined as an episode of acute sickle-cell-related pain requiring hospital assessment, analgesia, admission, or documentation as vaso-occlusive crisis in the clinical record. Acute chest syndrome was defined as a clinically diagnosed acute pulmonary complication in a woman with SCD, characterised by respiratory symptoms such as chest pain, cough, difficulty in breathing, hypoxia, or fever. Recurrent infections were defined as two or more clinically diagnosed infectious episodes during the index pregnancy, or documentation of repeated infection-related treatment or admission. Malaria was recorded separately when confirmed or clinically treated as malaria during pregnancy or admission.

Preeclampsia was defined as new-onset hypertension after 20 weeks of gestation, with systolic blood pressure of at least 140 mmHg and/or diastolic blood pressure of at least 90 mmHg, with proteinuria or another documented maternal organ dysfunction. Postpartum haemorrhage was defined as excessive bleeding after delivery documented by the attending clinical team, need for uterotonic treatment beyond routine prophylaxis, blood transfusion for post-delivery bleeding, surgical intervention for bleeding, or documentation of postpartum haemorrhage in the clinical record.

Acute kidney injury was defined as clinician-documented acute kidney injury, or an acute rise in serum creatinine above the local pregnancy reference range during pregnancy while deranged renal function was defined as any renal function test result outside the local laboratory reference range, including elevated serum creatinine, elevated urea, abnormal electrolytes judged clinically significant, or documentation of renal impairment. Deranged liver function was defined as any liver function test result outside the local laboratory reference range, including elevated transaminases, elevated bilirubin, or other clinically significant abnormality documented by the attending team.

Severe oligohydramnios was defined as ultrasound-documented markedly reduced amniotic fluid, including an amniotic fluid index below 5 cm, deepest vertical pocket below 2 cm, or clinical documentation of severe oligohydramnios. Antepartum haemorrhage was defined as vaginal bleeding after fetal viability and before delivery. Deep vein thrombosis and pulmonary embolism were recorded when clinically diagnosed, radiologically confirmed, or documented as such in the medical record. Chorioamnionitis was defined as clinical diagnosis of intra-amniotic infection based on maternal fever, uterine tenderness, fetal tachycardia, offensive liquor, or clinician documentation. Surgical-site infection or puerperal sepsis was defined as clinician-documented wound infection, puerperal sepsis, or postpartum infection requiring antibiotic treatment.

For fetal and neonatal outcomes, abortion was defined as pregnancy loss before fetal viability as documented in the clinical record. Stillbirth was defined as fetal death occurring at or after the gestational-age threshold for viability used in the study hospitals. Early neonatal death was defined as death of a liveborn baby before discharge or within the first seven days of life where this information was available. Low birthweight was defined as birthweight below 2500 g. Neonatal intensive care unit admission was recorded when the newborn was admitted to the neonatal unit for observation, treatment, or intensive care after delivery.

### Operational definition of HDU or ICU admission

During the study period, the two study hospitals implemented a local clinical protocol for pregnant women with SCD presenting in labour, with acute complications, or for delivery admission. Under this protocol, women were commonly placed initially in the high-dependency unit or intensive care unit for close maternal and fetal monitoring, early recognition of deterioration, and rapid escalation if complications developed. Therefore, in this study, HDU/ICU placement was defined as documented admission or placement in an HDU or ICU area during the pregnancy or delivery admission, irrespective of whether organ-support-level interventions were required.

This variable should therefore be interpreted primarily as a protocol-driven care-process indicator and marker of high-intensity observation within the local SCD pregnancy pathway. It should not be interpreted on its own as equivalent to organ-support-level critical illness, mechanical ventilation, vasopressor support, dialysis, or other forms of intensive organ support. For this reason, HDU/ICU placement was reported descriptively and was not used as a primary severe maternal morbidity outcome or as a predictor in the exploratory regression models.

### Sample size considerations

The study sample size was estimated for the second objective, which assessed adverse pregnancy outcomes among women with sickle cell disease (SCD). We used stillbirth as the sentinel adverse perinatal outcome because it is clinically important, consistently reported in comparable studies, and expected to occur more frequently among women with SCD than among women without SCD.

In a hospital-based study from India conducted between March 2011 and September 2015, SCD accounted for 131 of 10,519 delivery admissions (1.2%), while sickle cell trait accounted for 1,645 of 10,519 delivery admissions (15.6%). Stillbirth occurred in 9.9% of SCD delivery admissions compared with 4.2% among non-SCD delivery admissions, and low birthweight occurred in 70.2% of SCD deliveries compared with 43.8% among non-SCD deliveries (10). A retrospective study from Muhimbili National Hospital in Tanzania reported 149 SCD deliveries among 157,473 total deliveries from 1999 to 2011, equivalent to 95 SCD deliveries per 100,000 deliveries (4). In addition, unpublished routine data from Kawempe National Referral Hospital for financial year 2022/2023 showed 140 deliveries among women with SCD out of 25,100 total deliveries. Using OpenEpi for cohort studies based on the Fleiss method for rates and proportions, we assumed a two-sided 95% confidence level, 80% power, 4.2% stillbirth among unexposed/non-SCD delivery admissions, 9.9% stillbirth among exposed/SCD delivery admissions, and an unexposed-to-exposed ratio of 178.3, derived from the Kawempe National Referral Hospital delivery distribution. Under these assumptions, the minimum required number of exposed/SCD pregnancies was 129 using the Fleiss method and 146 after continuity correction. Allowing for 10% loss to follow-up or incomplete outcome ascertainment increased the target sample to 161 pregnant women with SCD.

The study therefore planned to recruit approximately 161 pregnant women with confirmed SCD from the two study sites using consecutive sampling. The final analytic cohort included 159 pregnant women with confirmed HbSS, which exceeded the uncorrected Fleiss minimum and was very close to the continuity-corrected target after allowance for incomplete follow-up. Because the study did not recruit a concurrent non-SCD comparison group, the sample size should be presented as supporting adequate precision and feasibility for descriptive estimation among women with SCD, while regression analyses should remain explicitly exploratory and hypothesis-generating.

### Data analysis

Analyses were primarily descriptive. Continuous variables were summarized as mean and standard deviation, with median and interquartile range presented where useful for describing skewed clinical variables. Categorical variables were summarized as n/N and percentage. Denominators exclude missing values and are shown explicitly. Multiple-response variables are presented as separate components; their percentages therefore do not necessarily sum to 100%. Cumulative incidence of maternal complications was estimated with 95% binomial confidence intervals.

### Regression modelling

For exploratory adjusted analyses, the preferred effect measure was the risk ratio. Log-binomial regression was attempted for rare outcomes, including maternal death and stillbirth, but these models did not converge and produced inadmissible fitted mean warnings. Therefore, modified Poisson regression with robust standard errors was retained as the primary risk-ratio approach across outcomes. Firth penalized logistic regression was retained only as a sparse-data sensitivity analysis for maternal death and stillbirth and is reported separately as odds ratios. Predictor selection considered statistical screening, clinical relevance, temporality, sparse-data limitations and avoidance of downstream delivery-management variables.

## Results

During the study period, 180 pregnant women with suspected or known SCD were screened at Kawempe National Referral Hospital and Mbale Regional Referral Hospital. Of these, 167 met the eligibility criteria and 161 provided written informed consent and were enrolled into the cohort. 2 women were excluded from the final analysis because pregnancy, delivery, or discharge outcome data could not be retrieved after review of available facility records and follow-up attempts. Based on these figures, the final mother-level analytic cohort was 159 women and 166 fetal outcomes as described in the STROBE participant flow in figure 1 below.

**Figure 1.**
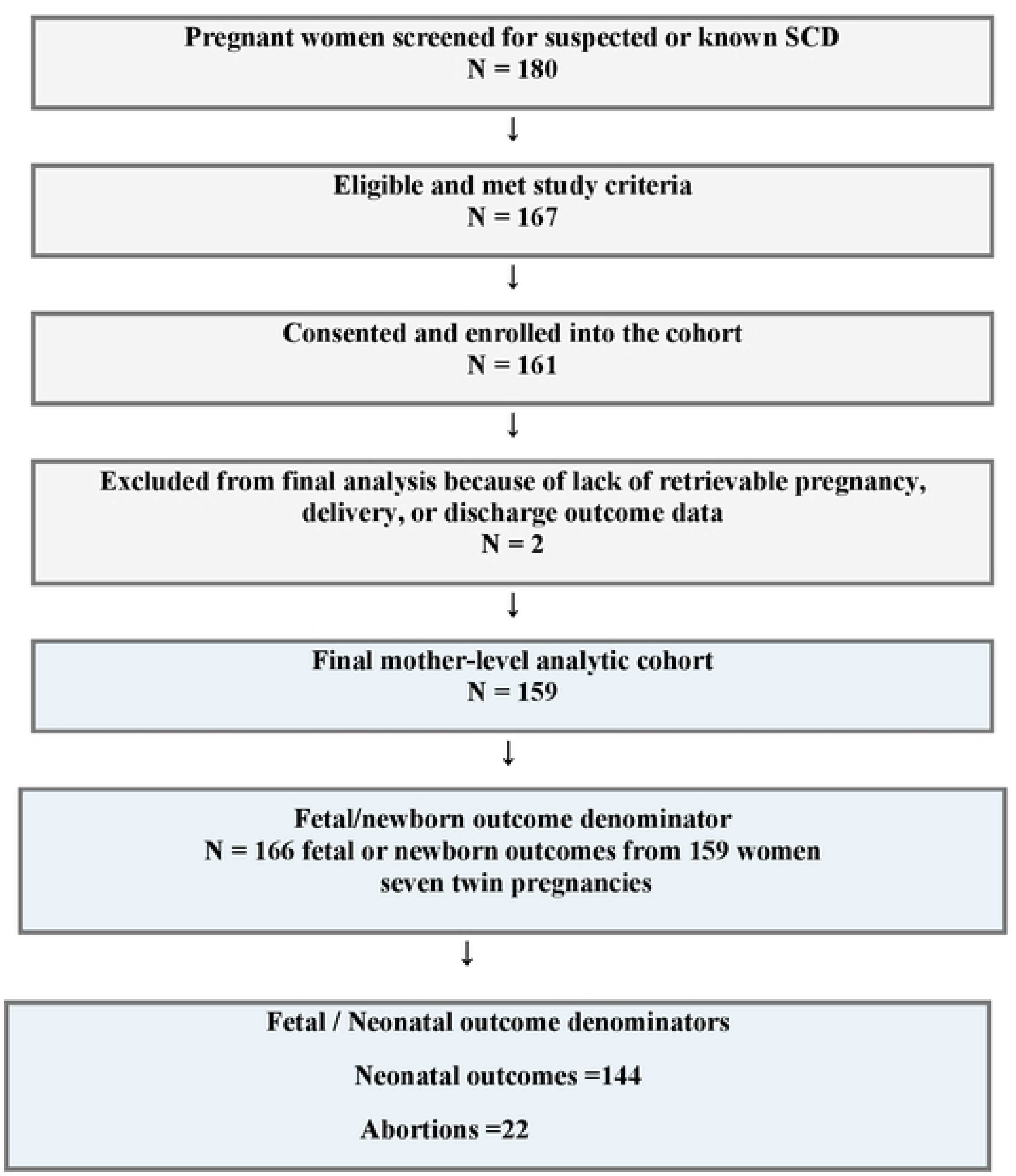
Participant flow for the SCD pregnancy cohort

Table 1 summarizes the sociodemographic, enrolment and obstetric profile of the 159 pregnant women with confirmed sickle cell disease that were included in the mother-level analytic dataset. The mean age was 24.3 years (SD 4.7), 109/159 (68.6%) were aged 20-29 years, and 82/159 (51.6%) were referred in rather than already known from the study-site antenatal clinic.

**Table 1.**
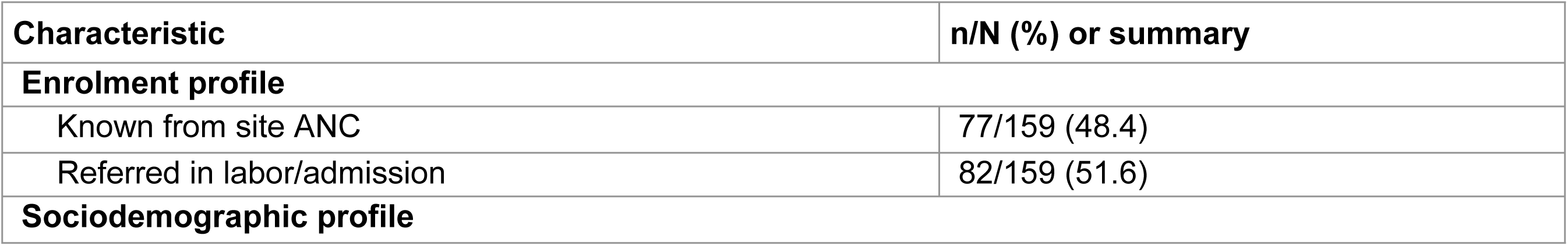
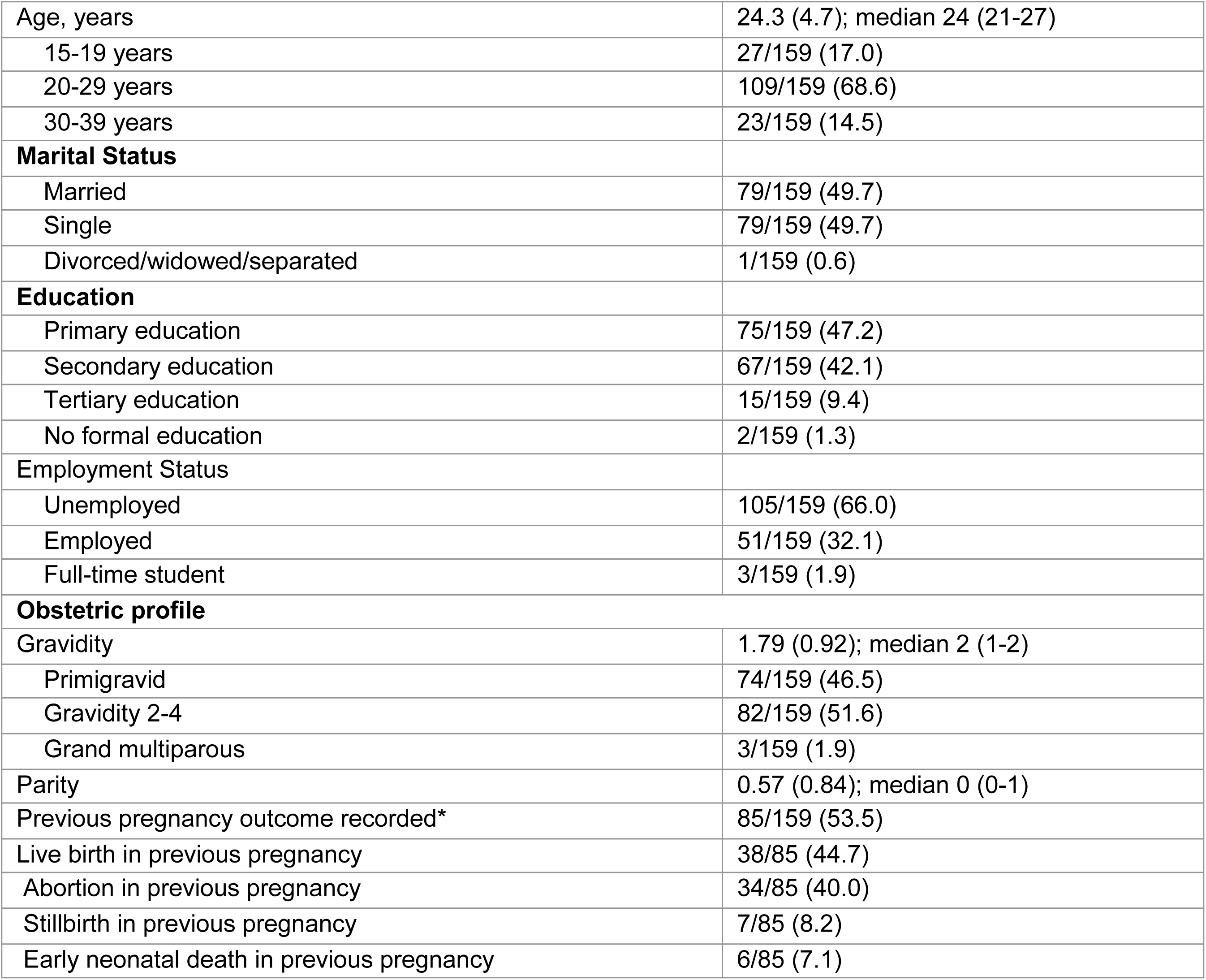
Sociodemographic, enrolment and obstetric profile of women with confirmed sickle cell disease.

| Characteristic | n/N (%) or summary |
| --- | --- |
| **Enrolment profile** |  |
| Known from site ANC | 77/159 (48.4) |
| Referred in labor/admission | 82/159 (51.6) |
| **Sociodemographic profile** |  |
| Age, years | 24.3 (4.7); median 24 (21-27) |
| 15-19 years | 27/159 (17.0) |
| 20-29 years | 109/159 (68.6) |
| 30-39 years | 23/159 (14.5) |
| **Marital Status** |  |
| Married | 79/159 (49.7) |
| Single | 79/159 (49.7) |
| Divorced/widowed/separated | 1/159 (0.6) |
| **Education** |  |
| Primary education | 75/159 (47.2) |
| Secondary education | 67/159 (42.1) |
| Tertiary education | 15/159 (9.4) |
| No formal education | 2/159 (1.3) |
| **Employment Status** |  |
| Unemployed | 105/159 (66.0) |
| Employed | 51/159 (32.1) |
| Full-time student | 3/159 (1.9) |
| **Obstetric profile** |  |
| Gravidity | 1.79 (0.92); median 2 (1-2) |
| Primigravid | 74/159 (46.5) |
| Gravidity 2-4 | 82/159 (51.6) |
| Grand multiparous | 3/159 (1.9) |
| Parity | 0.57 (0.84); median 0 (0-1) |
| Previous pregnancy outcome recorded* | 85/159 (53.5) |
| Live birth in previous pregnancy | 38/85 (44.7) |
| Abortion in previous pregnancy | 34/85 (40.0) |
| Stillbirth in previous pregnancy | 7/85 (8.2) |
| Early neonatal death in previous pregnancy | 6/85 (7.1) |

### Clinical, care and psychosocial profile during pregnancy

Table 2 summarizes clinical care and psychosocial characteristics of the participants which showed marked baseline clinical vulnerability. Majority 67/132(50.8%) had mild to moderate anaemia (Hb7.0-9.9 g/dl), High blood pressure (Systolic blood pressure > 140mmgh and or diastolic blood pressure >90mmgh) at admission was recorded in 9/159 (5·7%). Abnormal renal function test results were present in 114/148 (77·0%) and abnormal liver function tests in 116/148 (78·4%). Blood-transfusion history was common where 86/159 (54·1%) had received transfusion within the previous 3 months and 58/159 (36·5%) more than 3 months earlier. Hydroxyurea had been used before pregnancy by 77/159 (48·4%), but only 8/159 (5·0%) were recorded as currently using it during the pregnancy. Hospital use during pregnancy was substantial with 85/156 (54·5%) women having 1–2 admissions and 36/156 (23·1%) had 3–4 admissions. Antenatal-care attendance was mixed: 75/156 (48·1%) attended fewer than four visits, 77/156 (49·4%) attended 4–7 visits, and only 4/156 (2·6%) attended eight or more visits. Follow-up adherence was suboptimal, with 64/156 (41·0%) reporting attendance only sometimes and 17/156 (10·9%) rarely. Folic acid was the most recorded antenatal medication (141/155, 91·0%), whereas low-dose aspirin was received by 62/155 (40·0%) and clexane by 50/155 (32·3%).

**Table 2.**
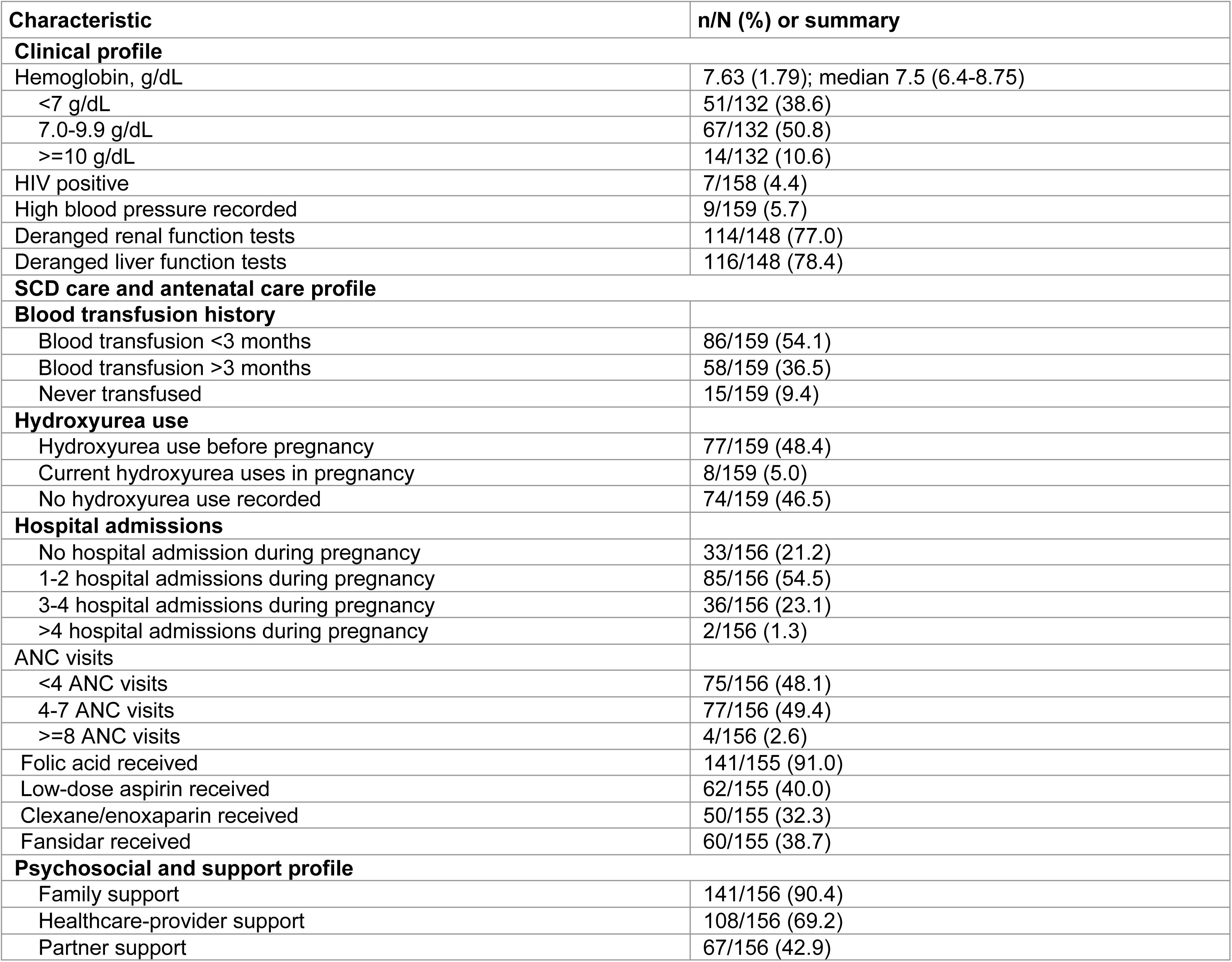
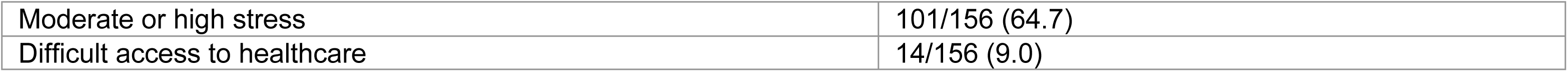
Clinical, care and psychosocial profile during pregnancy.

| Characteristic | n/N (%) or summary |
| --- | --- |
| **Clinical profile** |  |
| Hemoglobin, g/dL | 7.63 (1.79); median 7.5 (6.4-8.75) |
| <7 g/dL | 51/132 (38.6) |
| 7.0-9.9 g/dL | 67/132 (50.8) |
| >=10 g/dL | 14/132 (10.6) |
| HIV positive | 7/158 (4.4) |
| High blood pressure recorded | 9/159 (5.7) |
| Deranged renal function tests | 114/148 (77.0) |
| Deranged liver function tests | 116/148 (78.4) |
| **SCD care and antenatal care profile** |  |
| **Blood transfusion history** |  |
| Blood transfusion <3 months | 86/159 (54.1) |
| Blood transfusion >3 months | 58/159 (36.5) |
| Never transfused | 15/159 (9.4) |
| **Hydroxyurea use** |  |
| Hydroxyurea use before pregnancy | 77/159 (48.4) |
| Current hydroxyurea uses in pregnancy | 8/159 (5.0) |
| No hydroxyurea use recorded | 74/159 (46.5) |
| **Hospital admissions** |  |
| No hospital admission during pregnancy | 33/156 (21.2) |
| 1-2 hospital admissions during pregnancy | 85/156 (54.5) |
| 3-4 hospital admissions during pregnancy | 36/156 (23.1) |
| >4 hospital admissions during pregnancy | 2/156 (1.3) |
| **ANC visits** |  |
| <4 ANC visits | 75/156 (48.1) |
| 4-7 ANC visits | 77/156 (49.4) |
| >=8 ANC visits | 4/156 (2.6) |
| Folic acid received | 141/155 (91.0) |
| Low-dose aspirin received | 62/155 (40.0) |
| Clexane/enoxaparin received | 50/155 (32.3) |
| Fansidar received | 60/155 (38.7) |
| **Psychosocial and support profile** |  |
| Family support | 141/156 (90.4) |
| Healthcare-provider support | 108/156 (69.2) |
| Partner support | 67/156 (42.9) |
| Moderate or high stress | 101/156 (64.7) |
| Difficult access to healthcare | 14/156 (9.0) |

### Maternal Complications

Table 3 presents the maternal and intrapartum complications recorded among 159 pregnancies. Each complication was counted once per woman, even if repeated episodes occurred. The most frequently documented complications were anemia, reported in 93 pregnancies (58.5%, 95% CI 50.7–65.9), vaso-occlusive crisis in 87 pregnancies (54.7%, 95% CI 47.0–62.3) and Acute chest syndrome in 31 pregnancies (19.5%, 95% CI 14.1–26.3). Recurrent infections were recorded in 27 pregnancies (17.0%, 95% CI 11.9–23.6). Other common obstetric and maternal complications included postpartum hemorrhage in 21 pregnancies (13.2%, 95% CI 8.8–19.3) and preeclampsia in 17 pregnancies (10.7%, 95% CI 6.8–16.5). Less frequent complications included acute kidney injury, severe oligohydramnios, malaria, deep vein thrombosis, antepartum hemorrhage, pulmonary embolism, chorioamnionitis, and puerperal sepsis. No case of gestational diabetes was recorded. Intrauterine fetal death is included in the table because of its obstetric relevance, although it is a fetal rather than maternal complication.

**Table 3.** Women with at least one documented maternal or intrapartum complication during the index pregnancy, N=159.

| Complication | Events/N | Cumulative incidence, % (95% CI) |
| --- | --- | --- |
| Anemia | 93/159 | 58.5 (50.7-65.9) |
| Vaso-occlusive crisis | 87/159 | 54.7 (47.0-62.3) |
| Acute chest syndrome | 31/159 | 19.5 (14.1-26.3) |
| Recurrent infections | 27/159 | 17.0 (11.9-23.6) |
| Postpartum hemorrhage | 21/159 | 13.2 (8.8-19.3) |
| Preeclampsia | 17/159 | 10.7 (6.8-16.5) |
| Other complications | 16/159 | 10.1 (6.3-15.7) |
| Acute kidney injury | 8/159 | 5.0 (2.6-9.6) |
| Severe oligohydramnios | 7/159 | 4.4 (2.1-8.8) |
| Deep vein thrombosis | 5/159 | 3.1 (1.4-7.1) |
| Intrauterine fetal death* | 5/159 | 3.1 (1.4-7.1) |
| Malaria | 5/159 | 3.1 (1.4-7.1) |
| Chorioamnionitis | 4/159 | 2.5 (1.0-6.3) |
| Antepartum hemorrhage | 2/159 | 1.3 (0.3-4.5) |
| Pulmonary embolism | 1/159 | 0.6 (0.1-3.5) |
| Surgical-site infection/puerperal sepsis | 1/159 | 0.6 (0.1-3.5) |
| Gestational diabetes | 0/159 | 0.0 (0.0-2.4) |

\*Intrauterine fetal death is included because of its obstetric relevance but should be interpreted as a fetal outcome rather than a maternal complication.
Note: Percentages were calculated using 159 pregnancies as the denominator. Confidence intervals are Wilson 95% confidence intervals. Categories are not mutually exclusive; therefore, percentages do not sum to 100%.

### Maternal and fetal/newborn outcomes

Table 4 presents woman-level and pregnancy-level outcomes as fetal and newborn outcomes to avoid denominator ambiguity. Initial HDU/ICU placement was recorded in 145/159 women (91.2%). This high proportion reflects the local SCD pregnancy protocol, under which women presenting for labor, delivery admission, or acute complications were commonly placed in HDU/ICU areas for close monitoring and rapid escalation where needed. It should therefore be interpreted as a care-process indicator rather than as evidence that all women required organ-support-level critical care. Maternal death occurred in 15(9.4%) of the women and among 137 women with recorded mode of delivery, 107 (78.1%) had caesarean section while 30(21.9%) delivered vaginally. Of the 159 women in the final analytic cohort, 22 had abortions and were therefore included as pregnancy outcomes but excluded from delivery-mode analyses. Delivery-mode analysis was restricted to pregnancies that progressed to delivery after fetal viability and had documented mode of delivery. Denominators are shown explicitly for each outcome.

**Table 4.** Maternal, delivery, fetal and neonatal outcomes.

| Outcome | n/N (%) or summary |
| --- | --- |
| **Woman-level and pregnancy-level outcomes** |  |
| Initial HDU/ICU placement under local SCD protocol | 145/159 (91.2) |
| Maternal death | 15/159 (9.4) |
| Caesarean section among recorded deliveries | 107/137 (78.1) |
| Vaginal delivery among recorded deliveries | 30/137 (21.9) |
| Maternal length of stay after delivery, days | 5.7 (3.6); median 5 (4-7) |
| **Fetal/newborn outcomes as recorded in the mother-level file** |  |
| **Total number of fetuses =166** |  |
| Abortions (Pregnancy loss) | 22/166 (13.3) |
| Fetal/ Neonatal Outcomes | 144/166 (86.7) |
| Alive baby | 119 /144 (82.6) |
| Stillbirth | 25/144 (17.4) |
| Early neonatal death | 12/119 (10.1) |
| Birthweight, grams | 2194.5 (646.7); median 2220 (1800-2650) |
| <2500 g among recorded birthweights | 76/117 (65.0) |
| 2500-3499 g among recorded birthweights | 39/117 (33.3) |
| >=3500 g among recorded birthweights | 2/117 (1.7) |
| NICU admission among recorded newborns | 83/118 (70.3) |
| NICU length of stay, days | 4.0 (3.9); median 3 (2-5.5) |
| Alive at discharge among recorded newborns | 107/119 (89.9) |

*Denominators vary by outcome. Woman-level outcomes use the final analytic cohort of 159 women. Abortions were classified as pregnancy outcomes before delivery and were not included in delivery-mode denominators. Delivery-mode outcomes were restricted to women whose pregnancies progressed to delivery and had mode of delivery recorded. Fetal/newborn outcomes count each fetus or newborn separately, including twins. Early neonatal death was calculated among live births only.*

For fetal outcomes, the 159 women included (7) seven sets of twin pregnancies and therefore contributed 166 fetal outcomes. Among these 166 outcomes,22 (13.3%) were abortions while 119/ 144 (86.7%) resulted in a live birth but 12 (10.1 %) of these live births resulted in early neonatal deaths. Notably 25/144 (13.3%) were stillbirths while among newborns with APGAR recorded, 29/118 (24.6%) had a score below 7. Mean birthweight among newborns with recorded weight was 2194.5 g (SD 646.7), median 2220 g (IQR 1800-2650), and 76/117 (65.0%) weighed less than 2500 g. NICU admission was recorded for 83/118 newborns (70.3%); the most frequent indications were prematurity (70/83, 84.3%), low APGAR score (29/83, 34.9%), and low birthweight (24/83, 28.9%). Median NICU stay was 3 days (IQR 2-5.5).

### Exploratory models

Table 5 presents the primary exploratory adjusted models as risk ratios. Log-binomial models were attempted for rare outcomes but did not converge; modified Poisson models with robust standard errors were therefore used for the primary risk-ratio estimates. Firth logistic models are reported in the supplementary material as sensitivity analyses only. The regression findings should be interpreted cautiously because of small event numbers, missing data for some outcomes and the descriptive purpose of the study.

**Table 5.**
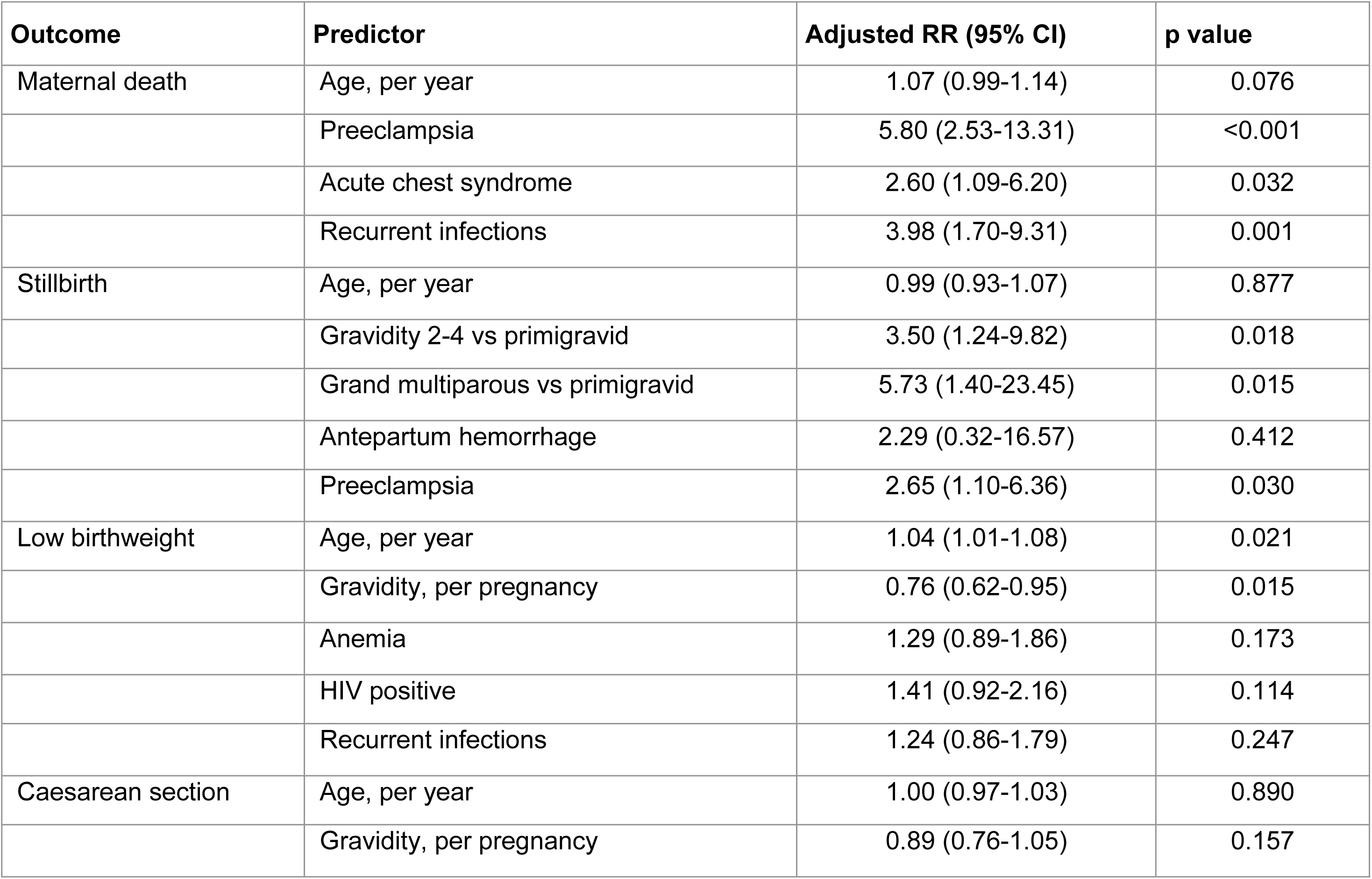
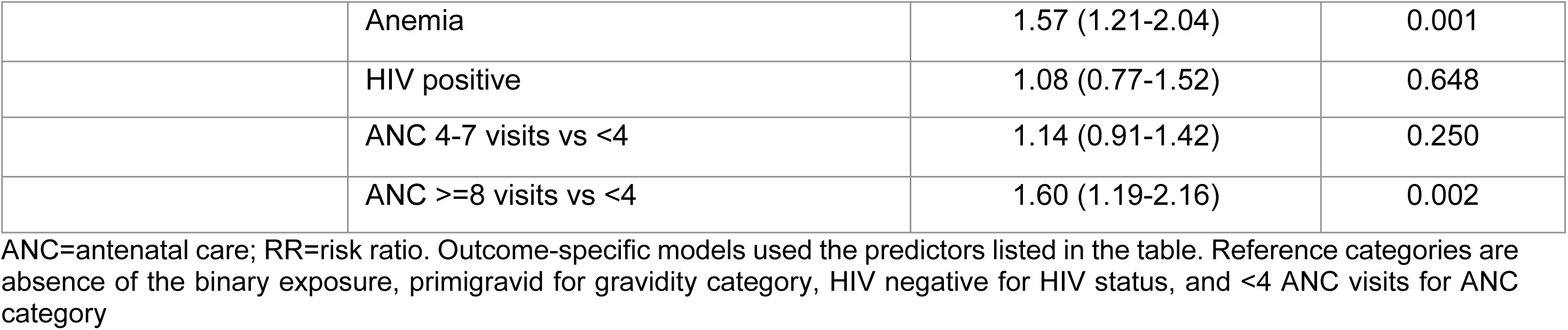
Primary exploratory adjusted associations with adverse outcomes.

## Discussion

In this prospective cohort of pregnant women with confirmed HbSS receiving care at two tertiary referral hospitals in Uganda, pregnancy was associated with a very high burden of maternal morbidity, substantial maternal mortality, and poor fetal and neonatal outcomes. The dominant clinical picture was one of severe baseline vulnerability, reflected by low haemoglobin concentrations, frequent recent transfusion, repeated hospital admission, and a high frequency of abnormal renal and liver function tests. The leading maternal complications were anaemia, vaso-occlusive crisis, acute chest syndrome, recurrent infections, postpartum haemorrhage, and preeclampsia. Fetal and neonatal outcomes were similarly adverse, with a high burden of pregnancy loss, stillbirth, early neonatal death, low birthweight, and neonatal-unit admission. Together, these findings reinforce the view that SCD pregnancy in referral-hospital practice is not a single-risk condition, but a multisystem high-risk state requiring coordinated obstetric, haematology, anaesthesia, transfusion, critical-care, and neonatal support(2,3,10).

The high frequency of maternal complications in this cohort is consistent with the established pathophysiology and clinical literature on SCD in pregnancy. Pregnancy increases circulatory demand, inflammatory activation, hypercoagulability, and oxygen requirements, while SCD is characterised by chronic haemolysis, vaso-occlusion, endothelial dysfunction, infection susceptibility, and progressive organ vulnerability. This combination plausibly explains the high frequency of anaemia, vaso-occlusive crisis, acute chest syndrome, infection, hypertensive disease, haemorrhage, and thromboembolic complications observed in this cohort. Systematic reviews and cohort studies have consistently shown that women with SCD have substantially higher risks of maternal morbidity, severe maternal morbidity, perinatal loss, preterm birth, fetal growth restriction, and neonatal complications than women without SCD (6,7,11–15).

The level of maternal mortality observed in this Ugandan cohort was particularly concerning. Although direct comparison across studies should be cautious because of differences in case mix, genotype, referral pathways, timing of enrolment, access to transfusion, and critical-care capacity, the proportion of maternal deaths in this cohort appears higher than in many specialist or mixed-setting cohorts. Studies from Tanzania and Ghana have shown increased maternal and perinatal risk among women with SCD, while work from Jamaica and Ghana suggests that maternal deaths can be reduced when care is delivered through structured multidisciplinary pathways with reliable surveillance, protocols, and escalation systems (4,5,16,17). These comparisons suggest that the excess risk observed in the present cohort may reflect not only biological severity, but also health-system factors, including late referral, reactive rather than preventive care, limited preconception optimisation, variable antenatal continuity, transfusion constraints, and delays in escalation.

The high proportion of women initially managed in HDU/ICU should be interpreted in the context of local practice. During the study period, the two hospitals used a local SCD pregnancy protocol under which women presenting in labour, for delivery admission, or with acute complications were commonly placed in HDU/ICU areas for close observation and rapid escalation. Therefore, HDU/ICU placement in this cohort is best understood as a protocol-driven care-process indicator and a marker of high-intensity monitoring, not as a direct measure of organ-support-level critical illness. This distinction is important because otherwise the 91.2% HDU/ICU placement could be misread as indicating that nearly all women required mechanical ventilation, vasopressor support, dialysis, or equivalent organ support. The finding nevertheless highlights the level of clinical vigilance required for SCD pregnancy in high-burden referral settings.

Several care-profile findings help explain the observed outcomes. More than half of the women were referred in labour or on admission rather than being already known from the study-site antenatal clinics, suggesting that many entered tertiary care late or during acute clinical need. Nearly half attended fewer than four antenatal-care visits, and very few reached eight or more visits. In SCD pregnancy, delayed presentation reduces opportunities for early risk stratification, optimisation of haemoglobin status, infection prevention and treatment, blood-pressure surveillance, fetal-growth monitoring, birth planning, and counselling on warning signs. Current guidance emphasises preconception counselling, early multidisciplinary antenatal review, serial fetal surveillance, planned delivery, access to transfusion support, thromboprophylaxis where indicated, and rapid response to acute chest syndrome, vaso-occlusive crisis, hypertensive disease, and infection(9,18–22)

The observed associations in the exploratory models are clinically plausible but should be interpreted cautiously. Maternal death was associated with preeclampsia, acute chest syndrome, and recurrent infections, each of which may represent a pathway to rapid maternal deterioration in SCD pregnancy. Preeclampsia may worsen endothelial dysfunction and placental disease; acute chest syndrome can lead to hypoxia and respiratory compromise; and recurrent infections may precipitate vaso-occlusion, anaemia, systemic inflammation, and multi-organ stress. The association between stillbirth and preeclampsia is also biologically plausible, given the role of placental insufficiency and maternal vascular disease in fetal compromise. The association between caesarean section and anaemia or higher antenatal-care attendance should not be interpreted as causal. It may reflect confounding by indication, whereby women recognised as clinically high risk or followed more closely were more likely to undergo operative delivery.

The perinatal findings show substantial fetal and neonatal vulnerability. Among pregnancies that progressed beyond abortion, the burden of stillbirth, early neonatal death, low birthweight, low APGAR score, and NICU admission was high. These outcomes are consistent with prior evidence that SCD pregnancy is associated with placental insufficiency, fetal growth restriction, preterm birth, perinatal loss, and increased neonatal-unit use(3,4,7,11,23–25). The low birthweight burden in this cohort was also broadly consistent with evidence from India, where Desai et al. reported a substantially higher proportion of low birthweight among SCD delivery admissions compared with non-SCD delivery admissions, alongside a higher stillbirth proportion among SCD deliveries(10). Although the Indian study differed in design, population, and denominators, it supports the wider observation that SCD pregnancy carries a high burden of fetal compromise across low-resource and middle-income settings.

The high caesarean-section proportion should also be interpreted carefully. In this cohort, caesarean section may partly reflect clinician concern about maternal or fetal compromise, including anaemia, fetal distress, previous obstetric risk, preeclampsia, and the need for controlled delivery in a high-risk setting. However, high operative-delivery rates also have implications for blood availability, anaesthetic readiness, postoperative monitoring, infection prevention, and future pregnancy risk. For women with SCD, caesarean delivery should therefore be embedded within a planned multidisciplinary pathway rather than treated as an isolated obstetric decision.

Overall, these findings support the need to strengthen SCD pregnancy care pathways in Uganda and similar high-burden African settings. Priority actions include early identification of girls and women with SCD before pregnancy, preconception counselling, early referral to specialist antenatal care, structured risk stratification at booking, reliable access to blood products, infection prevention and treatment, systematic blood-pressure and fetal-growth surveillance, planned delivery at facilities with obstetric, anaesthetic, transfusion, and neonatal capacity, and rapid escalation protocols for acute chest syndrome, severe anaemia, preeclampsia, sepsis, and haemorrhage. The findings also support stronger linkage between antenatal, delivery, postpartum, neonatal, and haematology services so that care is continuous rather than episodic.

**In conclusion**, pregnancy among women with SCD receiving tertiary referral care in Uganda was characterised by severe maternal morbidity, substantial maternal mortality, and poor fetal and neonatal outcomes. The results point to a need for proactive, structured, multidisciplinary SCD pregnancy care, with emphasis on early referral, preventive antenatal surveillance, transfusion readiness, rapid management of acute complications, and coordinated obstetric-neonatal care.

### Strengths and limitations

This prospective cohort study provides Uganda-specific evidence on pregnancy outcomes among women with confirmed HbSS receiving care at two tertiary referral hospitals. Its main strengths are the prospective design, genotype confirmation through the national reference laboratory, consecutive recruitment from two high-burden referral sites, explicit separation of woman-level and fetus/newborn-level denominators, and detailed reporting of both maternal complications and perinatal outcomes. These strengths are important because pregnancy-specific SCD data from Uganda remain limited, yet the country has a high burden of sickle cell disease and increasing survival of affected girls into reproductive age. However, the study had some limitations. The absence of a concurrent non-SCD comparison group limits direct estimation of excess risk relative to women without SCD in the same hospitals. In addition, there was no scheduled post-discharge follow-up, so late maternal and neonatal outcomes after discharge may have been missed.

The cohort was hospital based and likely enriched for severe referrals, so the findings may not represent all pregnant women with SCD in Uganda. Additionally, because women were enrolled at different gestational ages, including some at admission for delivery, the duration of prospective observation varied across participants and early pregnancy complications may have been under-ascertained for women who presented late. Notably, several variables had item-level missingness because some measurements were not done or were not documented in routine records; denominators are therefore reported explicitly. Finally, several regression outcomes were based on small numbers of events, so estimates, especially those with wide confidence intervals, should be interpreted cautiously and as hypothesis-generating rather than definitive.

## Ethics approval and consent to participate

The study was approved by the Makerere University School of Medicine Research and Ethics Committee (Mak-SOMREC-2023-805) and the Uganda National Council for Science and Technology (HS5315ES). Administrative clearance was also obtained from Kawempe National Referral Hospital and Mbale Regional Referral Hospital before participant recruitment. The study complied with the principles of the Belmont Report and the Declaration of Helsinki. Written informed consent was obtained from all participants before enrolment. Minors younger than 16 years were not included; participants aged 16-17 years were treated as emancipated minors and provided written informed consent. The consent process for participants aged 16–17 years as emancipated minors was approved by the ethics committees.

## Authors’ contributions

Conceptualisation: Jackline Akello, Sarah Kiguli, Annettee Nakimuli, David Mukunya, Ian Munabi, Musa Sekikubo, Grace Ndeezi.

Data curation: Jackline Akello, Joseph Rujumba, Grace Ndeezi, Kenneth Mugabe.

Formal analysis: Savio Mwaka, Joseph Rujumba, Jackline Akello, Annettee Nakimuli, Musa Sekikubo.

Methodology: Jackline Akello, David Mukunya, Ruth Namazzi, Pebalo Francis, Sarah Kiguli, Grace Ndeezi.

Resources: Sarah Kiguli, Ian Munabi, Ruth Namazzi, Grace Ndeezi.

Supervision: Musa Sekikubo, Annettee Nakimuli, Grace Ndeezi, Sarah Kiguli, Joseph Rujumba. Validation: Annettee Nakimuli, Joseph Rujumba, Sarah Kiguli, Grace Ndeezi.

Writing original draft: Jackline Akello, David Mukunya, Kenneth Mugabe, Savio Mwaka.

Writing review & editing: Jackline Akello, Sarah Kiguli, Grace Ndeezi, Musa Sekikubo, Savio Mwaka, Ian Munabi, Ruth Namazzi, Deogratias Munube, David Mukunya, Pebalo Francis, Kenneth Mugabe, Joseph Rujumba, Annettee Nakimuli.

## Availability of data and materials

The dataset contains potentially identifying and sensitive maternal and neonatal clinical information and cannot be made publicly available without restriction. De-identified data underlying the findings may be made available upon reasonable request to the corresponding author and with approval from the Makerere University School of Medicine Research and Ethics Committee and the Uganda National Council for Science and Technology, in accordance with the ethical approvals governing this study.

## Acknowledgements

We acknowledge Kawempe National Referral Hospital and Mbale Regional Referral Hospital for the administrative support provided during data collection. We also thank the ENRICH D43 administrative team for their support to the study. We are especially grateful to the women who participated at both study sites, whose contribution made this research possible. We further acknowledge Makerere University College of Health Sciences, particularly the Departments of Obstetrics and Gynaecology and of Pediatrics and Child Health, for their guidance, supervision, and institutional support.

